# Longitudinal effect of gestational age on the developmental trajectory of social competence difficulties from early childhood to mid-adolescence: Evidence from the UK Millennium Cohort

**DOI:** 10.1101/2020.06.09.20124537

**Authors:** Mariko Hosozawa, Noriko Cable, Yvonne Kelly, Amanda Sacker

**Author notes:** Correspondence to: Mariko Hosozawa, MD, PhD, Department of Epidemiology and Public Health, University College London, UK.

## Abstract

**Background:** Preterm birth and maternal psychological distress are associated with an increased risk for social competence difficulties; however, little is known about how this risk changes over time and across gestational age. This study aimed to examine mean developmental trajectories of social competence difficulties from early childhood to mid-adolescence by gestational age groups: very preterm (VP, <32 weeks), moderate-to-late preterm (MLP, 32-36 weeks), early-term (37-38 weeks) and full-term (39-41 weeks) and to assess the mediating effect of maternal psychological distress during infancy on the social competence difficulties trajectories.

**Methods:** Data were analysed on 15,821 participants from the UK Millennium Cohort Study participants, a nationally-representative birth cohort. Social competence difficulties were assessed by parent report when the participants were aged 3, 5, 7, 11 and 14 years. Maternal psychological distress was self-rated when the children were 9 months of age. Data were modelled using latent growth curve analysis.

**Results:** The developmental trajectories of social competence difficulties were u-shaped in all groups showing a decline from age 3 to 7, followed by a stable low period and then an increase during adolescence. VP children (n=173) showed pronounced difficulties throughout (b=0.94, SE=0.36 at age 14). MLP children (n=1,130) and early-term children (n=3,232) showed greater difficulties compared with their full-term peers around age 7, which resolved by age 14 (b=0.20, SE=0.13; 0.03, 0.07, respectively). Maternal psychological distress during infancy mediated 20% of the above association for VP.

**Conclusions:** The effect of gestational age on developmental trajectories of social competence difficulties can be dose-response. Monitoring and providing support on social development throughout childhood and into their adolescence and treating early maternal psychological distress may benefit preterm children, particularly those born VP.

---

Preterm birth (< 37 weeks completed gestation) is a known risk factor for children’s development, including cognitive, motor and behavioural problems (Schonhaut, Armijo, & Perez, 2015). Social competence is the ability to attain successful relationships with others in social situations (Rose-Krasnor, 1997), with difficulties associated with peer relationship problems (Ladd, 1999), educational underachievement (Henricsson & Rydell, 2006), internalizing and externalising behavioural problems (Huber, Plotner, & Schmitz, 2019), and poorer functioning in adulthood including increased risk of psychiatric problems (D. E. Jones, Greenberg, & Crowley, 2015), which are also elevated among children born preterm (Nosarti et al., 2012).

Growing evidence shows that children born very preterm (VP, <32 weeks) may be at increased risk for social competence difficulties across their lifespan compared with their full-term born peers (Eryigit-Madzwamuse, Strauss, Baumann, Bartmann, & Wolke, 2015; Healy et al., 2013; K. M. Jones, Champion, & Woodward, 2013; Ritchie, Bora, & Woodward, 2015). Some studies have also reported social competence difficulties in early childhood among children born moderate-to-late preterm (MLP, 32–36 weeks) (Cheong et al., 2017; S. Johnson et al., 2015; You, Yang, Hao, & Zheng, 2019), though with less consistency (Benzies, Magill-Evans, Ballantyne, & Kurilova, 2017; Heuser, Jaekel, & Wolke, 2018).

Despite this increased risk and the potential negative impact that social competence difficulties can have for preterm children, little is known about how the association between preterm birth and social competence difficulties changes dynamically across the course of development and how this varies by gestational age. Very few studies conducted a longitudinal, repeated assessment of social competence or its difficulties in the preterm population and most studies examined changes during early-childhood over short periods (i.e., up to age 8 and follow-up of fewer than 2 years) (Benzies et al., 2017; Heuser et al., 2018). One longitudinal study followed the trajectory of social difficulties beyond early-childhood among children born extremely preterm (<26 weeks) which found consistently greater difficulties compared with the full-terms peers (Linsell et al., 2019). However, the developmental course of social competence difficulties could differ by gestational age groups. That is, while the VP or extremely preterm children may continue to present social difficulties across development, preterm children born after a longer gestation (i.e., MLP children) could adapt and ‘catch-up’ to their full-term peers as they age, as shown in studies of cognitive development (Fitzpatrick, Carter, & Quigley, 2016). Also, despite emerging evidence showing higher risks of developmental problems among those born at early-term (37-38 weeks) compared to the full-term born children, evidence regarding social development of this group is scarce. Taken together, it is important to understand the developmental course of social competence difficulties from childhood and into adolescence across a more fine-grained range of gestational age groups; this information could inform us ‘who’ and ‘when’ to target to improve the social outcomes of preterm children.

The mechanism underlying increased social competence difficulties among children born preterm remain largely unknown. Studies hypothesise that in addition to the biological effect of prematurity and associated treatment stress, postnatal environmental factors that influence early parent-child interaction may also play a role (Healy et al., 2013; Montagna & Nosarti, 2016). Preterm birth could be a stressful experience for the parents, particularly mothers, leading to increased psychological distress during their child’s infancy (Carson, Redshaw, Gray, & Quigley, 2015; Vigod, Villegas, Dennis, & Ross, 2010) which could affect early parent-child interaction (Korja et al., 2008) and subsequent social development. Therefore, maternal psychological distress during infancy could be a modifiable mediator on the pathway between preterm birth and later social competence difficulties. However, the potentially mediating effect of maternal psychological distress has not been examined in relation to social competence difficulties across gestational age groups. Parenting style may also influence the child’s social development (Kennedy & Bakeman, 1984); however, we did not find a significant association between gestational age groups and parenting styles measured at 9 months or aged 3 in this study, therefore, the mediating effect of parenting style was not included in our model.

Using a nationally-representative population-based cohort in the United Kingdom (UK), this study examines developmental trajectories of social competence difficulties from early childhood to mid-adolescence by gestational age groups (i.e., VP, MLP, early-term and full-term). We also assess the role of maternal psychological distress during infancy in the association between gestational age and social competence difficulties. We focused on maternal psychological distress during infancy as mothers of preterm children are more likely to experience psychological distress during their child’s infancy (Carson et al., 2015) and this period is known as a sensitive period for children’s development (Bagner, Pettit, Lewinsohn, & Seeley, 2010). We hypothesise that trajectories of social competence difficulties will differ by gestational age group, with the VP group showing consistent difficulties over time while the negative risks among MLP and early-term groups will become smaller over time. We also hypothesise that early maternal psychological distress will mediate the observed associations.

## Methods

### Study sample

The Millennium Cohort Study (MCS) is a population-representative cohort study which follows the health and development of children born in the UK between September 2000 and January 2002 (Hansen, 2014). Data were collected when the cohort members were 9 months and 3, 5, 7, 11, 14 years old. The study used a stratified cluster design to oversample children living in disadvantaged areas and those with a high proportion of ethnic minority groups; further details of the study design are described elsewhere (Hansen, 2014). The MCS is approved by an NHS Research Ethics Committee and written consent was obtained from all participating parents at each survey (Hansen, 2014). The use of anonymised data for academic purposes did not require additional ethical approval. Of the 18,818 children who participated at the first wave of the study (i.e., age 9 months), we excluded those whose main respondent was not the natural mother (n = 57), those whose gestational age or birth weight was missing (n = 210) or implausible for birth weight, defined as birth weight lying outside of ± 4 SD based on gender-segregated birth weight for gestational age centile charts (n =134). Those who were born post-term (≥ 42 weeks, n = 665) were also excluded due to known poor agreement between maternal recall and hospital records for this group in the MCS (Poulsen et al., 2011). This left 17,752 children. In this study, children with valid information on their social competence difficulties on at least one of all five waves (i.e., age 3, 5, 7, 11 and 14 interviews) were included, resulting in 15,821 children in the analytic sample.

### Gestational age groups

Gestational age in weeks was calculated based on the mother’s report of the expected due date, which corresponded well with gestational age recorded in the linked hospital data (Poulsen et al., 2011). Children were divided into four groups based on their reported- gestational age:

- VP (23–31 weeks completed gestation)
- MLP (32–36 weeks)
- Early-term (37–38 weeks)
- Full-term (39–41 weeks).

### Social competence difficulties score

Social competence difficulties during childhood and adolescence were measured using peer and prosocial subscales of the parent-rated Strengths and Difficulties Questionnaire (SDQ)(Goodman, 1997), measured at ages 3, 5, 7, 11 and 14. Peer and prosocial subscales of the SDQ represented the two key concepts of social competence (i.e., prosocial behaviour representing the appropriateness of the behaviours in social situations, and social initiation/ peer competence representing active participation in social interactions; (Rydell, Hagekull, & Bohlin, 1997). The total of peer and reverse-coded prosocial scale scores were summed to give a social competence difficulties score (range 0-20, higher scores indicate more severe difficulties). In our sample, Cronbach’s alpha for social competence difficulties ranged from 0.65 to 0.74 between waves indicating acceptable internal consistency.

### Maternal psychological distress

Maternal psychological distress was assessed using a modified version of the Rutter Malaise Inventory measured when the child was approximately 9 months old (Johnson, Atkinson, & Rosenberg, 2015). This is a 9-item self-completed questionnaire asking about emotional disturbance and associated physical symptoms. It has good reliability and correlates well with the original 24-item version that has good validity (J. Johnson et al., 2015). The scores range from 0 to 9 with higher scores indicating more severe psychological distress.

### Covariates

The following covariates, all measured at 9 months, were included in the study model as potential confounders: child’s sex, multiple birth indicator, highest maternal education attainment (attaining GCSE Advanced level or equivalent which is the qualification level required to enter university, or not), relative income poverty of the household (indicated by household equivalised income of less than 60% of the UK national median household income (Hansen, 2014), and mother’s age at the cohort children’s birth.

### Data Analyses

We first conducted a descriptive analysis to examine the association between the exposure and the study variables. We also conducted a sample bias analysis to explore whether children in our analytic sample were different from those not in the analytic sample on our study variables.

To predict the children’s social competence difficulties over time by gestational age groups, we modelled the group-average trajectory of social competence difficulties using latent growth curve models. We included linear and quadric slopes to account for the curved shape of children’s average trajectories. Chronological age was used as the time scores to take into account the considerable variation in the children’s age at each interview. Here age was centred at age 14 (i.e., the last available interview conducted) to explore group differences at that age and all the models were adjusted for sex and multiple birth status. In Model 0, intercept, linear and quadric slope factors were included. In Model 1, gestational age was included to demonstrate the association between gestational age and the child’s social competence difficulties at age 14 and how it changed with age. Model 2 further controlled for time-invariant family confounders (i.e., maternal education, household income, maternal age at childbirth, measured at 9 months). We examined possible mediation of the association by maternal psychological distress when the child was 9 months old in two ways: 1) adjusting this factor in the model (Model 3) and 2) examining the effect in a mediation model. Further, to aid our understanding of the group difference at each age point, we repeated the analyses by centring at the beginning of the study period (i.e., age 3) and around the midpoint (i.e., age 7). Moderation in associations by sex and by maternal psychological distress in the growth curve models was examined using an interaction term (i.e., gestational age*sex or gestational age*maternal psychological distress); however, there was little evidence of effect modification by sex or by maternal psychological distress. As a sensitivity analysis, we repeated the analyses by restricting the sample to those with a valid social competence score at all five time-points (n = 10,460). Growth curve models and mediation analysis were conducted using Mplus 8.3. (Muthén & Muthén, Los Angeles, CA) and descriptive analyses and figure plotting were conducted using Stata version 15.1 (Stata Corp, College Station, TX). All analyses took into account the clustering and stratified sampling of the MCS and were weighted using survey weights.

### Missing data

Missing data on covariates ranged from 0.1% on maternal education to 3.4% on maternal psychological distress. To minimize data loss, we imputed missing covariates using multiple imputation by chained equations using all the variables included in the analysis model plus some auxiliary variables to improve the imputation. Regression analyses were run across 20 imputed datasets and adjusted using Rubin’s rules (White, Royston, & Wood, 2011). Imputed results were broadly similar to those obtained using observed samples and therefore the imputed results are presented here; see Table S1 for characteristics of the observed data.

## Results

Descriptive characteristics of our study sample by gestational age groups are presented in Table 1. Of the 15,821 children included, 11,286 (71.2%) were born full-term, 3,232 (20.4%) as early-term, 1,130 (7.3%) as MLP and 173 (1.1%) as VP. Compared with the full-term group, multiple births were more frequent among all the preterm groups with 22.7% of children in the VP group born with multiple birth status. Mothers of the early-term and MLP had lower educational attainment compared with the full-term group, however, the groups did not differ on the proportion with low household income. Mean maternal psychological distress when the child was 9 months was significantly higher in the VP group compared to the other groups (mean score 2.4 for VP vs 1.6–1.7 for other groups). The result of our sample bias analysis showed that those from more disadvantaged families and with younger mothers were more likely to be excluded from the analysis. However, there was no difference in the distribution of gestational age groups or mean maternal psychological distress (Table S2). The mean score for social competence difficulties at each age by gestational age groups are presented in Table S3.

**Table 1.** Descriptive characteristics of the multiply imputed sample, by gestational age groups

|  | Full-term<br>(n =11,288,<br>71.2%) | Early-term<br>(n = 3,232,<br>20.5%) | MLP<br>(n =1,131,<br>7.3%) | VP<br>(n = 173,<br>1.1%) | P<sup>a</sup> |
| --- | --- | --- | --- | --- | --- |
|  | % | % | % | % |  |
| Mean gestational age, wk<br>(SE) | 40.2 (0.0) | 37.9 (0.0) | 35.1 (0.0) | 29.1 (0.2) | < .001 |
| Mean birth weight, kg<br>(SE) | 3.5 (0.0) | 3.2 (0.0) | 2.5 (0.0) | 1.3 (0.0) | < .001 |
| Sex (Male) | 50.5 | 52.5 | 54.4 | 54.7 | .06 |
| Multiple birth | 0.3 | 5.3 | 18.6 | 22.7 | < .001 |
| Low maternal education | 49.5 | 53.9 | 55.6 | 52.9 | < .001 |
| Low household income | 26.7 | 28.0 | 28.0 | 29.3 | .53 |
| Maternal age at<br>childbirth, mean (SE) | 29.0 (0.1) | 29.4 (0.2) | 29.4 (0.3) | 28.5 (0.6) | < .001 |
| Maternal psychological<br>distress, mean (SE)<sup>b</sup> | 1.6 (0.0) | 1.7 (0.0) | 1.7 (0.1) | 2.4 (0.2) | < .001 |

*Note:* Weighted means and percentages are shown. MLP=moderate-to-late preterm; VP=very preterm; SE=standard error. <sup>a</sup>P values for group difference. <sup>b</sup>Measured when the child was 9 months.

The estimated results for the growth curve models examining the association between gestational age and social competence difficulties from age 3 to 14 are shown in Table 2 (estimates with the full set of covariates are shown in Table S4). In Model 1, the coefficient increased from full-term to lower gestational age group, with the VP group showing significantly different associations with social competence difficulties at age 14. The coefficient for the interaction between gestational age group and linear and quadric slopes were insignificant indicating that the overall change in trajectory was similar across groups. Adjusting for family confounders in Model 2 did not change the result. Restricting our analysis to children with social competence difficulties score for all five-time points showed similar results (Table S5).

**Table 2.**
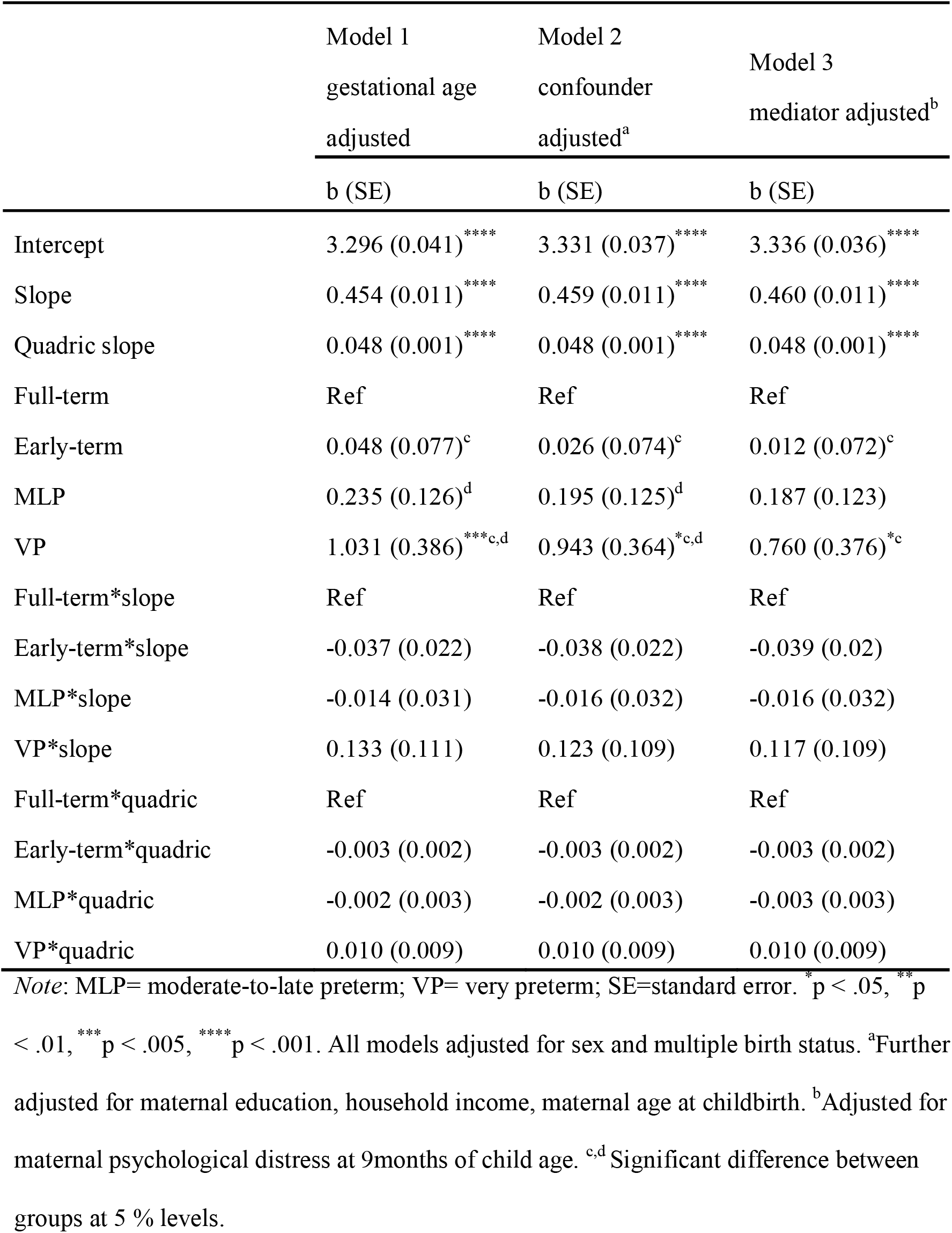
Fixed estimates of latent growth curve models of social competence difficulties from age 3 to 14

Figure 1 shows the trajectories plot predicted from Model 2. The mean trajectory of social competence difficulties was u-shaped for all gestational age groups; declining from age 3 to around age 7, followed by a stable low period and then increasing from around age 11 to age 14. The VP group showed a consistently elevated trajectory compared to all the other groups from age 3 to 14 suggesting higher levels of social competence difficulties throughout childhood and into mid-adolescence. Despite no evidence for an interaction between gestational age group and linear and quadric slopes, a small difference for the MLP group from that of the full-term group emerged around age 5 (i.e., the age when children start school in the UK) and persisted throughout mid-childhood then gradually narrowed during adolescence, resulting in no significant difference from the full-term groups at age 14. The figure also suggests that the trajectory for the early-term group overlapped with that of the MLP group but with a smaller gap from the full-term group during mid-childhood; the difference from the full-term group was no longer evident at around age 14. These unique group differences at each age were further supported by our supplementary analyses centred at different ages; while only the VP group showed a significant difference from the full-term group at age 3, all groups (i.e., early-term, MLP and VP groups) showed a significant difference from the full-term group at age 7 (Table S6).

**Figure 1.**
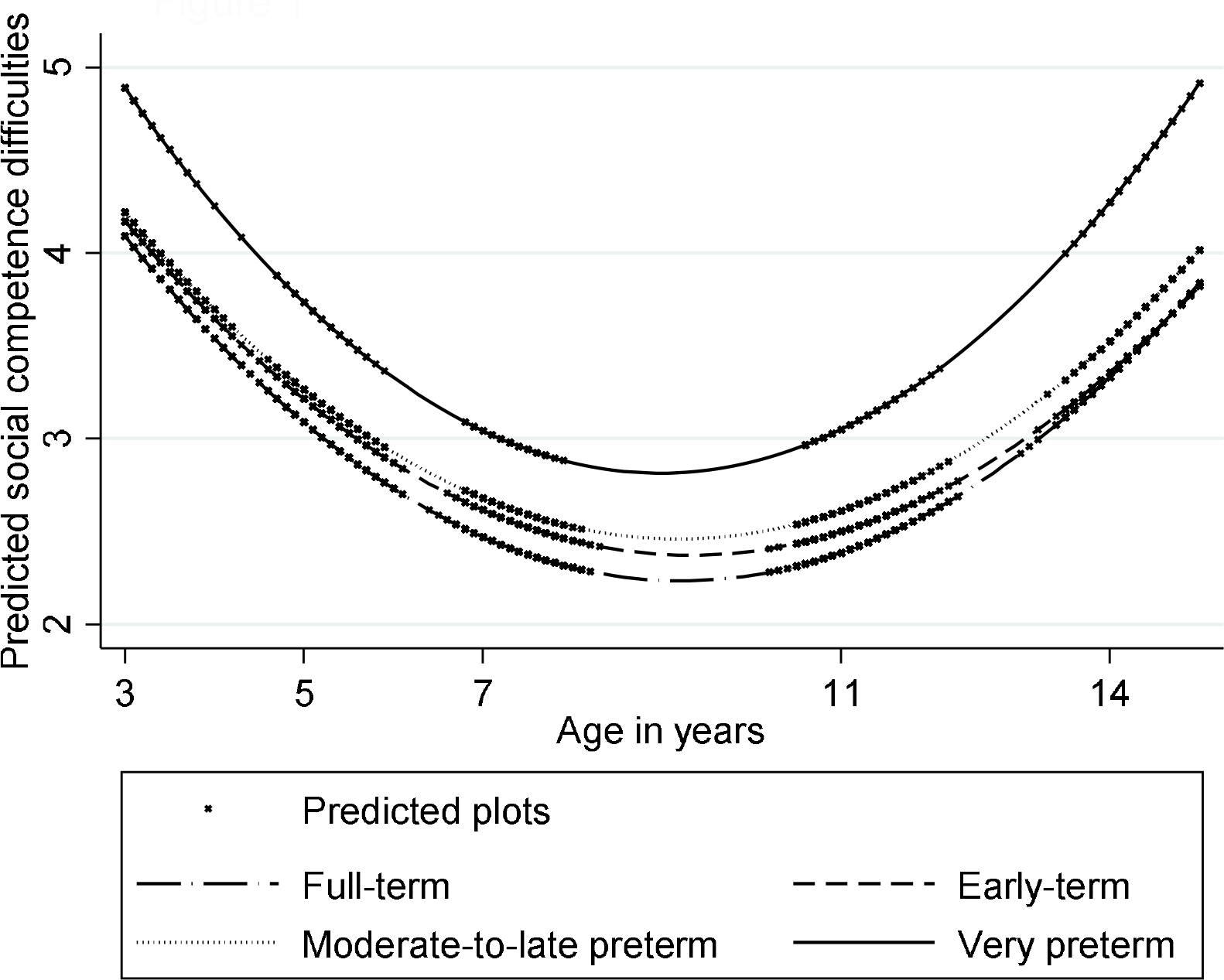
Predicted plot and trajectory of social competence difficulties by gestational age groups from the confounder adjusted model (Model 2).

In model 3, maternal psychological distress was added, showing that it was related to levels of social competencies but not to changes in social competence scores, mainly attenuating the very preterm birth and social competence difficulties coefficients (Table 2). Mediation analyses (Table 3, full estimates are presented in Table S7) confirmed that maternal psychological distress partially mediated the association between very preterm birth and the social competence difficulties intercept, accounting for 20% of the association.

**Table 3.** Estimates of direct and indirect effect for intercepts (mediated through maternal psychological distress) of association between gestational age groups and social competence difficulties.

|  | Indirect effect | Direct effect | Total effect |
| --- | --- | --- | --- |
| Effect size | b (SE) | b (SE) | b (SE) |
| Full-term | Ref | Ref | Ref |
| Early-term | 0.022 (0.010)* | 0.012 (0.072) | 0.034 (0.074) |
| MLP | 0.024 (0.015) | 0.187 (0.123) | 0.211 (0.124) |
| VP | 0.188(0.041)**** | 0.760 (0.376)* | 0.949 (0.366)* |

*Note:* MLP=moderate-to-late preterm; VP=very preterm; SE=standard error. \* p < .05, \*\*\*\* p < .001. Mediating effect for the intercept of latent growth curve model in Mplus adjusted for sex, multiple birth status, maternal education, household income and maternal age at childbirth are shown.

## Discussion

This study aimed to identify the trajectories of social competence difficulties across the full-range of gestational age groups from early childhood and into adolescence, using a nationally representative cohort in the UK. The trajectories of social competence difficulties were u- shaped across gestational age groups; declining during early childhood to around age 7, followed by a stable low period and then escalating as the children entered adolescence. While the VP group showed pronounced difficulties throughout, the MLP and early-term groups showed slightly greater difficulties around age 7 compared with the full-term group which each resolved by age 14. Maternal psychological distress when the child was 9 months old partially mediated the association between gestational age groups and social competence difficulties for VP.

Our results support our hypothesis that trajectories of social competence difficulties would differ by gestational age group. Our findings that VP children showed persistently greater social competence difficulties from early-childhood into mid-adolescence concurred with previous studies (Eryigit-Madzwamuse et al., 2015; Healy et al., 2013; K. M. Jones et al., 2013; Ritchie et al., 2015). Trajectory analyses demonstrated that these difficulties for the VP children did not decrease with age compared with the full-term children. One may wonder how clinically meaningful these significant between-group differences may be. In our study, the size of the coefficient difference from the full-term group for the VP group at age 14 was approximately 0.32 SD of the population average score at that age. Albeit using a different measure, a previous study demonstrated that a 1 SD increase in their social competence scores measured at age 5 was associated with various adulthood functioning including 66% increased odds of having stable employment or 46% reduction in the number of years on medication for mental health conditions (D. E. Jones et al., 2015). Taken together, persistently greater difficulties across childhood resulting in 0.32 SD difference at age 14 for the VP group could have a prolonged effect on their future social and emotional functioning. The MLP and early-term groups showed greater difficulties compared with the full-term children at around age 7, however, the gap was no longer significant at age 14. This finding may partly contrast with studies reporting higher social competence difficulties among children born MLP(Cheong et al., 2017; S. Johnson et al., 2015; You et al., 2019). The difference in findings could be due to different sampling method (population-based in our study vs hospital-based (Cheong et al., 2017; S. Johnson et al., 2015; You et al., 2019) or the difference in measured abilities as some studies measured a wider social-emotional competence (Cheong et al., 2017; S. Johnson et al., 2015; You et al., 2019). As the previous studies were conducted during early-childhood, our result may suggest that if MLP children are followed-up into adolescence, their social competence difficulties may decline to a similar level as their full-term peers. Furthermore, despite finding different associations for point estimates of the coefficients at different ages for MLP and early-term groups, the interaction between gestational age groups and linear or quadric slopes was insignificant, therefore, it would be important in the future to replicate our result in larger samples to confirm whether the suspected trajectories for the MLP and early-term groups are supported. In our study, maternal psychological distress during infancy partially mediated the association between very preterm birth and social competence difficulties. Possible explanations for this could be that early psychological distress among parents of preterm children may affect parental perceptions of the child (e.g., regard the child as more vulnerable) (Silverstein, Feinberg, Young, & Sauder, 2010); parenting behaviours such as less responsive or overprotective parenting (Gerstein, Njoroge, Paul, Smyser, & Rogers, 2019; Silverstein et al., 2010) and parent-child interactions (Korja et al., 2008), which may all impact the child’s subsequent social development. Given that mothers of preterm children, especially very preterm children, are vulnerable to psychological distress during the early years (Carson et al., 2015; Vigod et al., 2010), our results highlight the importance of interventions aiming to reduce maternal psychological distress during infancy, particularly among mothers of the VP children. This approach may reduce the long-term social competence difficulties of very preterm children.

Interestingly, the developmental trajectory of social competence difficulties was u-shaped. Our results correspond with studies exploring the developmental trajectory of social competence in the general population which suggest an inverted u-shape trajectory (i.e., increase during early-childhood (Santos, Vaughn, Peceguina, Daniel, & Shin, 2014), stability during mid-childhood (Cote, Tremblay, Nagin, Zoccolillo, & Vitaro, 2002), and a decline during adolescence (Luengo Kanacri, Pastorelli, Eisenberg, Zuffiano, & Caprara, 2013). Further, as these prior studies suggest that social competence may rebound and increase in late adolescence towards early-adulthood (Luengo Kanacri et al., 2013), future studies extending the observation to adulthood would elaborate how the gap in social competence difficulties across gestational groups observed in this study could continue to evolve.

### Strengths and Limitations

Our study has many strengths, including the use of nationally-representative cohort data in the contemporary UK, which enabled us to investigate the association between finer-grained gestational age groups and social competence difficulties. The longitudinal design of the MCS with repeated measurements of the child’s social competence difficulties over five waves allowed us to identify the curvilinear u-shaped trajectories across childhood and into mid-adolescence.

There are some limitations to our study. First, our social competence difficulties were measured based on parental report. While parental-report is valuable in identifying children’s behavioural difficulties, it could have biased the results by reflecting parental views or expectations (Dickey & Blumberg, 2004). Employing multi-informant assessment and/or including objective measurement of social competence and its difficulties may be useful (Sekigawa-Hosozawa, Tanaka, Shimizu, Nakano, & Kitazawa, 2017). Second, although we used a population-representative sample, children from disadvantaged families were more likely to be excluded from our analysis. This could have underestimated our findings because low socioeconomic position is associated with both preterm birth and social competence difficulties. Third, in our mediation analysis, the child’s social competence difficulties and maternal psychological distress were measured mostly by the same person (i.e., the mother). This could have inflated the estimates as distressed mothers are more likely than non-distressed mothers to report negatively of their child’s behaviour (Silverstein et al., 2010). However, there was a 13-year time-lag between the measurement of our mediator and increased psychological distress during infancy among mothers of very preterm children are reported to resolve with time (Treyvaud, 2014). There is also a possibility that the observed association between preterm birth, level of maternal psychological distress and child’s social competence difficulties was confounded by a shared common genetic factor. However, currently, evidence regarding a shared genetic liability is inconclusive (Jansen et al., 2018; Leppert et al., 2019).

## Conclusion

Our result suggests a dose-response effect of gestational age on developmental trajectories of social competence difficulties and may indicate the following clinical implications. Given the increased social competence difficulties across development, it is important to monitor aspects of social development and provide continuous support into adolescence for children born VP. Providing social skill training to the child and interventions aiming to reduce maternal psychological distress during infancy may benefit these preterm children at risk.

Further research is needed to understand how social competence developmental trajectories evolve into adulthood, and to explore the lasting impact of increased social competence difficulties not only for children born VP, but also children born MLP and early-term. This information would help improve the long-term care for these children.

## Data Availability

Data were obtained via the UK Data Archive. Further information about the study is found at https://cls.ucl.ac.uk/cls-studies/millennium-cohort-study/.

Key Points

- Children born preterm are at increased risk of social competence difficulties, how this risk changes as they develop and across the full-range of gestational age remains unknown.
- The trajectories of social competence difficulties were u-shaped across groups and the very preterm (VP) group showed pronounced difficulties throughout childhood and into mid-adolescence.
- Compared with full-terms, moderate-to-late preterm (MLP) children and early-term born children showed greater difficulties around age 7, however, this resolved by age 14.
- Maternal psychological distress during infancy partially mediated the above association for VP.
- Continuous monitoring and support into adolescence on social development and treating early maternal psychological distress may particularly benefit children born VP.

## Acknowledgement

This work was supported by the UK Economic and Social Research Council (ES/R008930/1) and the Japan Foundation for Paediatric Research. The authors thank the Millennium Cohort Study families for their time and cooperation, as well as the Centre for Longitudinal Studies (CLS) and the UK Data Service for the use of data. None of the founders, CLS or the UK Data Service were involved in conducting this study and holds no responsibility for the analysis or interpretation of the data.

